# Critical Arterial Stenosis Revisited

**DOI:** 10.1101/2022.04.24.22274211

**Authors:** Joseph P Archie

## Abstract

**Introduction:** Stenosis of an organ/tissue primary artery can produce ischemia or only reduce blood flow reserve. Despite incomplete hemodynamic understanding of critical arterial stenosis, degree of diameter stenosis continues to be an index for patient management. This study aims to use the law of conservation of energy to quantitate the arterial pressure gradient produced by stenosis, determine organ/tissue perfusion pressure, blood flow and reserve as a function of degree of diameter stenosis and determine ischemic critical diameter stenosis.

**Methods:** The three-component model is parallel stenotic artery and collateral arteries supplying an organ/tissue. The three hemodynamic variables are blood pressure, blood flow and frictional percent diameter stenosis. Two new non-dimensional variables, K and C, are introduced to simplify understanding. K is the magnitude of arterial diameter stenosis produced energy dissipation. C is the magnitude of potential collateral blood flow. Conservation of energy analysis of arterial stenosis gives the pressure gradient produced and stenosis vascular resistance. Organs/tissues intrinsically autoregulate blood flow when perfusion pressure is greater than threshold value. Model energy analysis defines collateral vascular resistance and gives perfusion pressure, blood flow and reserve as a function of diameter stenosis. Results are illustrated in both pressure-stenosis domain and blood flow-stenosis domain. Renal and internal carotid artery (ICA) stenosis numerical results are in cm/gram/sec (CGS) units.

**Results:** The magnitude of arterial stenosis energy dissipation is proportional to K, a fourth power function of stenosis diameter with a steep slope between 65% and 80%, (K = 67 to 625). Organs/tissues without collaterals, C = 0, have specific critical arterial stenosis values within this range. For a renal artery with average diameter and blood flow critical stenosis is 74% (K = 233). Organ tissues with collateral blood flow potential equal to their normal resting blood flow have C = 1.0. Those with poor collaterals, C < 1, have critical stenosis from 65% to 99% depending on collateral magnitude, 0 < C < 1. In this group critical ischemic ICA stenosis begins at 70% and up to 99%, (K = 132 to ∞). Organ/tissues with good collateral circulation, C > 1, do not have ischemic critical stenosis, including the ICA. However, in these patients stenosis progression reduces blood flow reserve.

**Conclusion:** Arterial stenosis may or may not produce an ischemic critical value for specific organ/tissue supplied depending on the magnitude of collateral circulation.

## Introduction

Based on intraoperative blood flow measurements we reported in 1981 that critical internal carotid artery (ICA) diameter stenosis, was approximately 60%^1^. Several discussants correctly pointed out that the relationship between blood flow and pressure make quantification of critical arterial stenosis difficult and one considered critical arterial stenosis “ pursuit of an impossible goal”. Our 60% value intuitively seemed low. In retrospect, the study failed because collateral blood flow reduced measured ICA flow and the calculated critical stenosis value. Decades later the critical stenosis problem remains inadequately analyzed as does the effect of arterial stenosis on organ/tissue hemodynamics in general. One might expect success using modern computational fluid dynamics (CFD) to numerically solve complex fluid mechanics problems that includes stenosis turbulent flow, but to date this is not the case^2,3^. The observation that smaller pipe diameter produces an increase in pressure drop (pressure gradient) in proportion to the lumen area was made by Poiseuille centuries ago. Much later It was found that decreasing stenosis area in animal arteries increases pressure gradient^4,5^. Some investigators recognized the value of using conservation of energy to capture stenosis turbulence^6,7^ but did not consider critical stenosis in the context of recipient organ/tissue. Standard CFD based on the law of conservation of momentum, the Navier-Stokes equations including Reynolds stresses and/or empirical functions, incompletely quantitates turbulent flow. Conversely, the law of conservation of energy encompasses turbulence and provides the potential to determine the effect of arterial stenosis on supplied body organ/tissues. Recent studies demonstrate that stenosis produced turbulent kinetic energy (TKE) can be quantitively measured by magnetic resonance imaging (MRI), is not reversible back to pressure energy and dissipated locally^8,9^. Directly measured aortic valve and vessel phantom stenosis pressure gradients correlate well with MRI measured TKE dissipation^10^. This information plus the necessity to consider arterial stenosis in the context of specific artery-collateral-organ/tissue systems and energy analysis offers an opportunity for determining critical arterial stenosis. This study aims to use the law of conservation of energy to; quantitate the arterial pressure gradient produced by stenosis, determine organ/tissue perfusion pressure, blood flow and reserve as a function of degree of diameter stenosis and identify critical diameter arterial stenosis, if it exists.

## Methods

### Hemodynamic Energy

Energy generated by the left ventricle and delivered to systemic organs/tissues is essentially all pressure (≈ 99% pressure energy and ≈ 1% kinetic energy). In normal body systems, blood flow is laminar and hemodynamic energy is primarily dissipated efficiently in body organs/tissues by parallel small artery branches, arterioles and pre-capillaries. Conversely, arterial stenosis converts pressure energy to TKE that is dissipated, reducing organ/tissue perfusion pressure. The hemodynamics of turbulent blood flow is poorly understood and incompletely modeled by both CFD^3^ and classical fluid dynamics^11^. Prior clinical studies of critical arterial stenosis using laminar flow or empirical approximations also fail^12,13^. Conservation of energy includes turbulence, circumventing the problem, an opportunity to succeed.

### Assumptions and Definitions

Blood is assumed to be an incompressible, homogenous and isotropic fluid. Volume blood flows, Q, and pressures, P, are mean values. Systemic blood pressure, P_a_, is arterial pressure minus venous pressure, a normalization. The artery has cylindrical lumen area A (diameter D) with a concentric stenosis of lumen area a (diameter d). System hemodynamic input pressure energy is P_a_Q, and the 1% input kinetic energy is neglected. Vascular resistance, R, a defined calculated variable, is pressure gradient,_Δ_P, divided by flow, Q, or_Δ_P = RQ. Pressure energy is Q_Δ_P = RQ^2^ and energy density is_Δ_P = RQ. A simple three-component energy model is analyzed; stenotic artery anatomically in parallel with collaterals supplying organ/tissue blood flow. Hemodynamic energy is dissipated by the vascular resistances; stenosis, R_s_, collateral, R_c_, and organ/tissue, R_o_. Fractional percent diameter stenosis (F%DS, X) is the independent variable.

Numerical calculations are centimeter, gram, second (CGS) units. Pressure is mmHg (1mmHg = 1,333 dyne/cm^2^), blood flow is cm/sec, energy is dyne cm/sec (erg/sec) = 10^−7^ watts and blood density is 0.994 g/ml.

### Arterial Stenosis Energy Dissipated and Vascular Resistance, R_s_

Conservation of mass requires that volume blood flow (Q = mean velocity V times area A) in the normal artery segment equal that in the stenosis lumen, AV_1_ = aV_2_ and Q = AV_1_ = aV_2._ Conservation of energy requires that stenosis produced kinetic energy equal the decrease in pressure energy. For a straight uniform diameter artery with a focal stenosis the transfer of pressure energy to kinetic energy is (P_1_ - P_2_)Q_s_ = Q_s_ (V_2_^2_^ V_1_^2^)/2, where Q_s_ is blood flow, A is normal lumen area, V is normal resting mean blood flow velocity and location 1 is upstream from a stenosis at location 2. Assuming a concentric round stenosis with lumen area A (diameter D) and stenosis area a (diameter d) the pressure gradient is ΔP = (P_1_ - P_2_) = Q_s_^2^[(1/a)^2^ – (1/A)^2^]. The NASCET definition of percent diameter stenosis is 100(1 – d/D) and fractional percent diameter stenosis, X = (1 – d/D). Lumen areas are, a =_π_d^2^/4 and A =_π_D^2^/4. The stenosis generated kinetic energy density is ΔP = Q_s_^2^{[1/(1 – X)^4^] – 1}/2A^2^, or ΔP = Q_s_^2^K/2A^2^, where Q_s_ is artery blood flow. The kernel K (term that contains the variable X) is K = {[1/(1 – X)^4^] – 1}, a universal measure of the magnitude of stenosis produced kinetic energy, independent of Q_s_. This is given in Figure 2 as K versus X. Arterial stenosis K values between 67 and 625 correspond to fractional percent diameter stenosis, X, values from 65% to 80% diameter stenosis (slope of curve approximately 37:1). The wide variance in stenosis produced kinetic energy is associated with a narrow range of degree of stenosis. Over 98% of stenosis produced kinetic energy is turbulent, irreversible back to pressure energy and dissipated in the stenosis zone. Vascular stenosis resistance, defined as the ratio of pressure gradient to flow, is R = ΔP/Q. Arterial stenosis vascular resistance is R_s_ = Q_s_{[1/(1 – X)^4^] – 1}/2A^2^ = Q_s_K/2A^2^, where the subscript_s_ indicates arterial stenosis resistance (R_s_) and blood flow (Q_s_). This result, like K, stands-alone and is universally applicable to body systems.

### Organ/tissue Vascular Resistance, R_o_

Organ/tissues control blood flow by intrinsically autoregulating perfusion pressure energy to meet metabolic energy requirements. The lower autoregulation threshold perfusion pressure (P_cut_) means that organs/tissues have two blood flow zones depending on perfusion pressure. Lassen’s measurements of normal human cerebral blood flow autoregulation^14,15^ with an ischemic perfusion pressure threshold of 50mmHg is illustrated in Figure 1. Renal autoregulation lower threshold for normal glomerular filtration is ≈ 80mmHg and for ischemia is ≈ 60mmHg. In the autoregulation zone, vascular resistance is R_o_ = P_p_/Q_n_, where Q_n_ is normal blood flow and P_p_ is perfusion pressure. If organ/tissue P_p_ falls below autoregulation pressure threshold, P_cut_, blood flow is linearly pressure dependent, R_o_ = P_cut_/Q_n_, and Q < Q_n_. That is, P_p_ = (P_cut_/Q_n_)Q.

**Figure 1.**
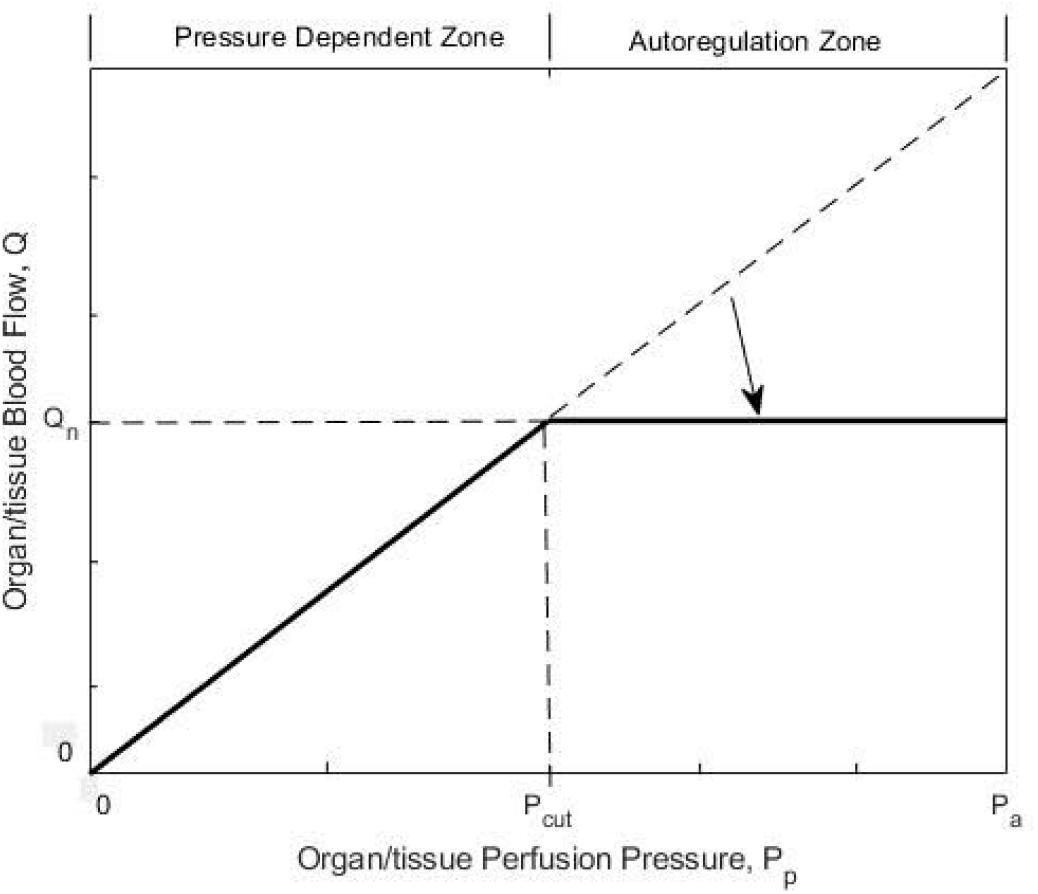
Schematic of Lassen cerebral blood flow autoregulation when organ/tissue perfusion pressure, P_p_, is greater than pressure threshold, P_cut_. Normal blood flow is Q_n_.

### Collateral Vascular Resistance, R_c_

Many organs/tissues have both a primary artery and collaterals. However, kidneys are essentially entirely dependent on renal artery blood flow and accessory renin arteries supply isolated kidney segments. Brain has a complex blood supply with both primary arteries and collaterals primarily the circle of Willis. Collateral vascular resistance, R_c_, is individual and organ/tissue specific, and of potential magnitude C. This is a result of energy conservation analysis of the three-component system.

At the organ/tissue autoregulation-perfusion pressure zones interface and artery occlusion, X = 1.0, the energy density is P_a_ = (R_c_ + R_o_)Q_n_, or R_c_ = (P_a_/Q_n_ - R_o_), where R_o_ can take either the autoregulation zone or the pressure dependent zone value. Choosing the latter, R_o_ = P_cut_/Q_n_, collateral vascular resistance is R_c_ = (P_a_ - P_cut_)/Q_n_. This meets the requirement that R_c_ be a constant. To make this specific for systems and individuals R_c_ = (1/C)(P_a_ - P_cut_)/Q_n_, where C the magnitude of collateral blood flow potential and ranges from zero to infinity. When C = 0, R_c_ = ∞ (no collaterals) and when C = ∞, R_c_ = 0. When K = ∞, the artery is occluded and organ/tissue perfusion pressure is due to collateral flow and clinically referred to as stump or back pressure, P_stump_^16^. The relationship between the magnitude of collateral vascular resistance, C, to P_stump_ is C = (P_stump_/P_cut_)(P_a_ - P_cut_)/(P_a_ - P_stump_). These are consistent with the three-component system’s pressure and blood flow end-points, C from 0 to ∞ and P_stump_ from 0 to P_a_.

### Three Component Hemodynamic Energy

The parallel stenotic artery and collateral pressure total energy supplied to organ/tissue is (P_a_ - P_p_)Q = ΔPQ = R_t_Q^2^, where R_t_ =(R_s_R_c_)/(R_s_ + R_c_). Organ/tissue received energy is R_o_Q^2^ (energy density R_o_Q) where R_o_ = P_p_/Q_n_ in theautoregulation perfusion pressure zone and R_o_ = P_cut_/Q_n_ in the lower perfusion pressure dependent zone. Artery vascular resistance R_s_ contains F%DS (X), the independent variable.

Solutions are organ/tissue perfusion pressure and blood flow as functions of percent diameter stenosis and include critical arterial stenosis values if they exist. Critical stenosis, X = X_crit_, occurs if stenotic and collateral energy dissipation equals organ/tissue lower autoregulation perfusion pressure energy threshold.

## Results

### Stenosis Energy Dissipation and Vascular Resistance

Dissipated stenosis energy density is ΔP = Q_s_^2^K/2A^2^, where Q_s_ is artery blood flow and K = {[1/(1 – X)^4^] – 1}, a universal measure of the magnitude of arterial stenosis produced TKE. Figure 2 illustrates K versus X, (F%DS). While the wide K variance for a narrow range of degree of stenosis is a stand-alone result, it is key to organ/tissue systems analysis. Arterial stenosis vascular resistance is R_s_ = Q_s_K/2A^2^.

**Figure 2.**
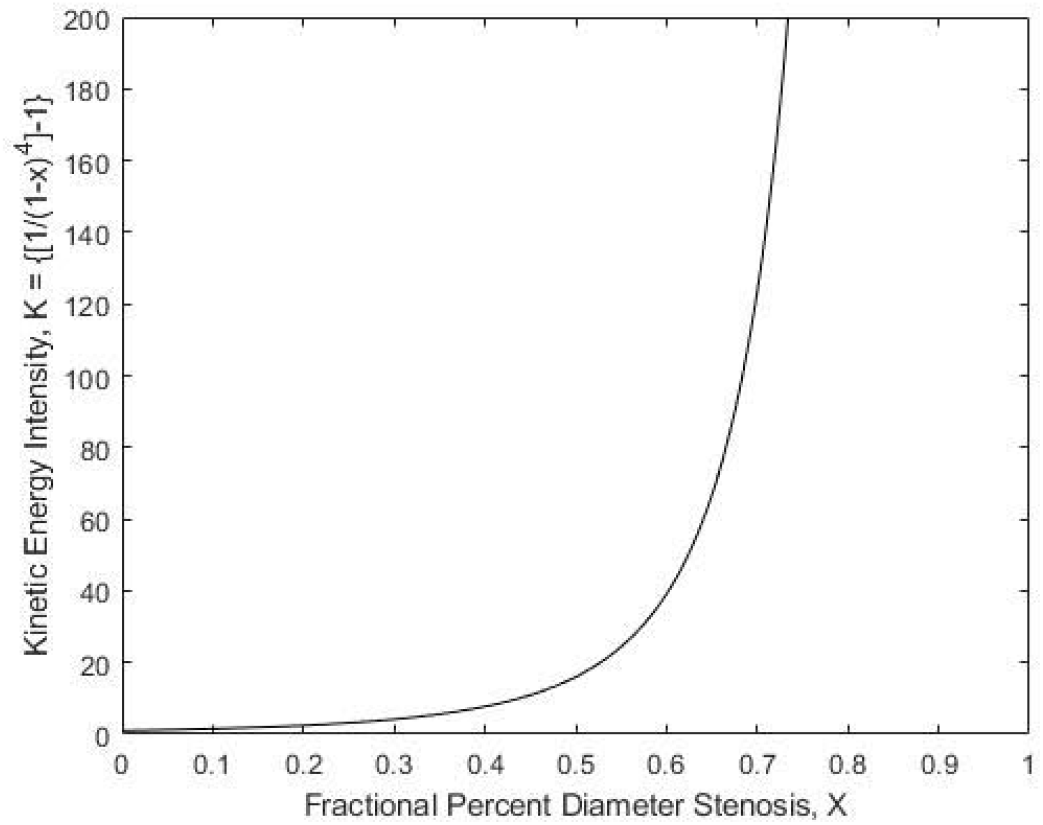
Arterial stenosis produced magnitude of dissipated energy, K, as function of fractional percent diameter stenosis, X.

### Collateral Vascular Energy and Resistance

This is a constant, specific for each organ/tissue system and individuals. At the organ/tissue autoregulation-perfusion pressure zones interface and artery occlusion, X = 1.0, the energy density is P_a_ = (R_c_ + R_o_)Q_n_, or R_c_ = (P_a_/Q_n_ - R_o_), where R_o_ can take either the autoregulation zone or the pressure dependent zone value. For the latter, R_o_ = P_cut_/Q_n_ and collateral vascular resistance is R_c_ = (P_a_ - P_cut_)/Q_n_. This meets the requirement that R_c_ be a constant. To make this specific for systems and individuals R_c_ = (1/C)(P_a_ - P_cut_)/Q_n_, where C, a measure of the magnitude of collateral blood flow potential and ranges from zero to infinity. When C = 0, R_c_ = ∞ (no collaterals) and when C = ∞, R_c_ = 0. When K = ∞, the artery is occluded and organ/tissue perfusion pressure is due to collateral flow and clinically referred to as stump or back pressure, P_stump_^16^. The relationship between the magnitude of collateral vascular resistance, C, to P_stump_ is C = (P_stump_/P_cut_)(P_a_ - P_cut_)/(P_a_ - P_stump_). This is consistent with the three-component system’s pressure and blood flow end-points, C from 0 to ∞ and P_stump_ from 0 to P_a_.

### Organ/tissue Perfusion Pressure and Diameter Stenosis

Conservation of energy for the three component system is P_a_Q = (R_t_ + R_o_)Q^2^. When organ/tissue perfusion pressure P_p_ is equal to or higher than P_cut_,R_o_ =P_p_/Q_n_ and Q=Q_n_, giving P_p_ =P_a_ -Q_n_R_t_. With R_s_ and R_c_ the result is P_p_ = P_a_ – Q_n_/[2A^2^/Q_s_K + (P_a_ - P_cut_)/C Q_n_]. The functional relationships between organ/tissue perfusion pressure energy density, P_p_, and fractional percent diameter stenosis, X, is given in Figure 3 when C = 0 (no collaterals), and in Figure 4 for the kidney data (no collaterals) and Figure 5 for the cerebral hemisphere (no collaterals) examples. In Figure 5 the perfusion pressure curves in the autoregulation zone (P_p_ > P_cut_) illustrates the decrease in reserve energy density and blood flow reserve as stenosis progresses and collateral magnitudes C = 0, 0.33, 1.0 and 2.0.

**Figure 3.**
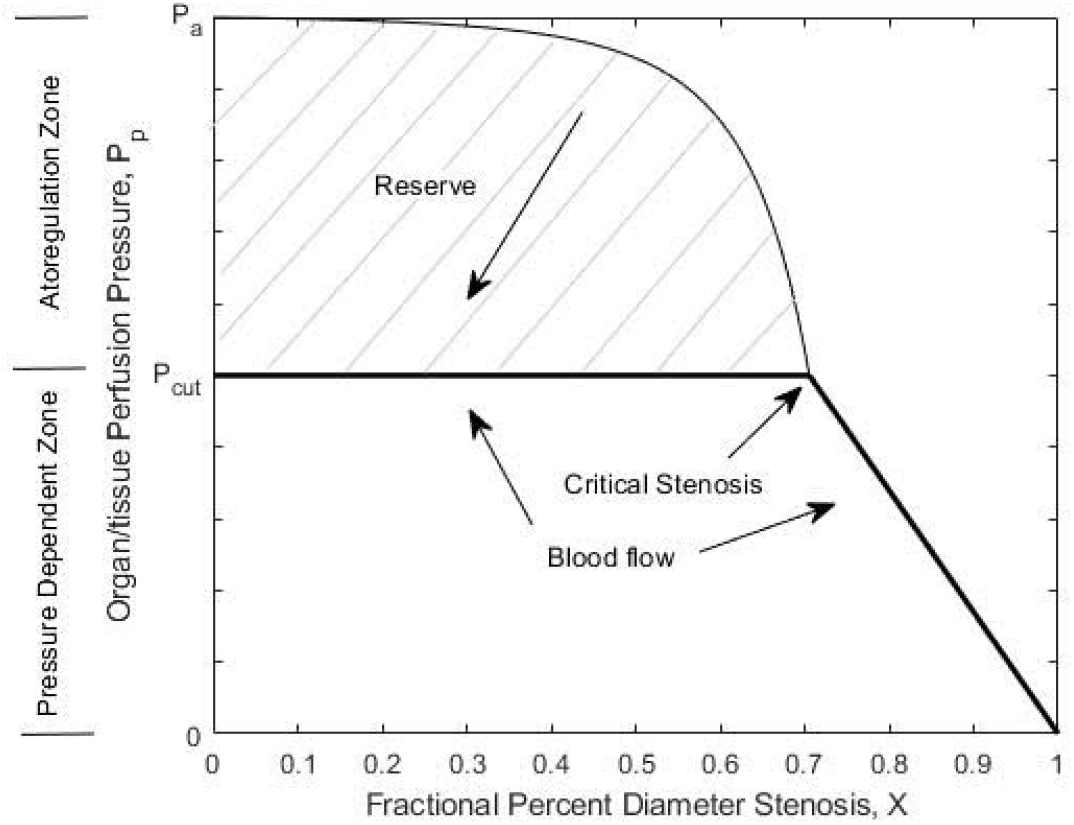
Energy solution for organ/tissue perfusion pressure and blood flow versus fractional percent diameter stenosis, X, in the absence of collaterals. In the upper pressure-stenosis domain stenosis is critical when P_p_ reaches P_cut_. The blood flow heavy lines are blood flow-stenosis domain where Q = Q_n_ up to critical stenosis and the ischemic blood flow at higher X values is slightly non-linear.

**Figure 4.**
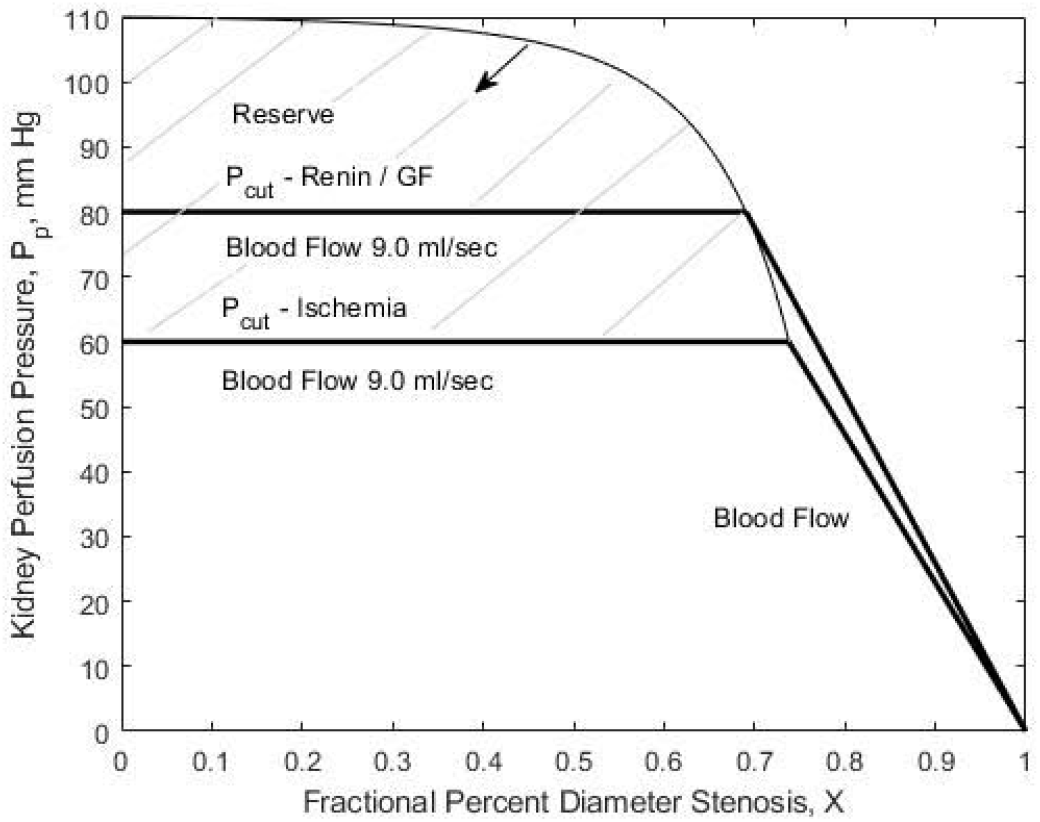
Numerical solution for kidney with two critical stenosis values, ischemia (74%) and glomerular filtration (71%). Blood flow has heavy lines are in the flow-stenosis domain. The renin dysfunction critical value is probably closer to 40%-50%.

**Figure 5.**
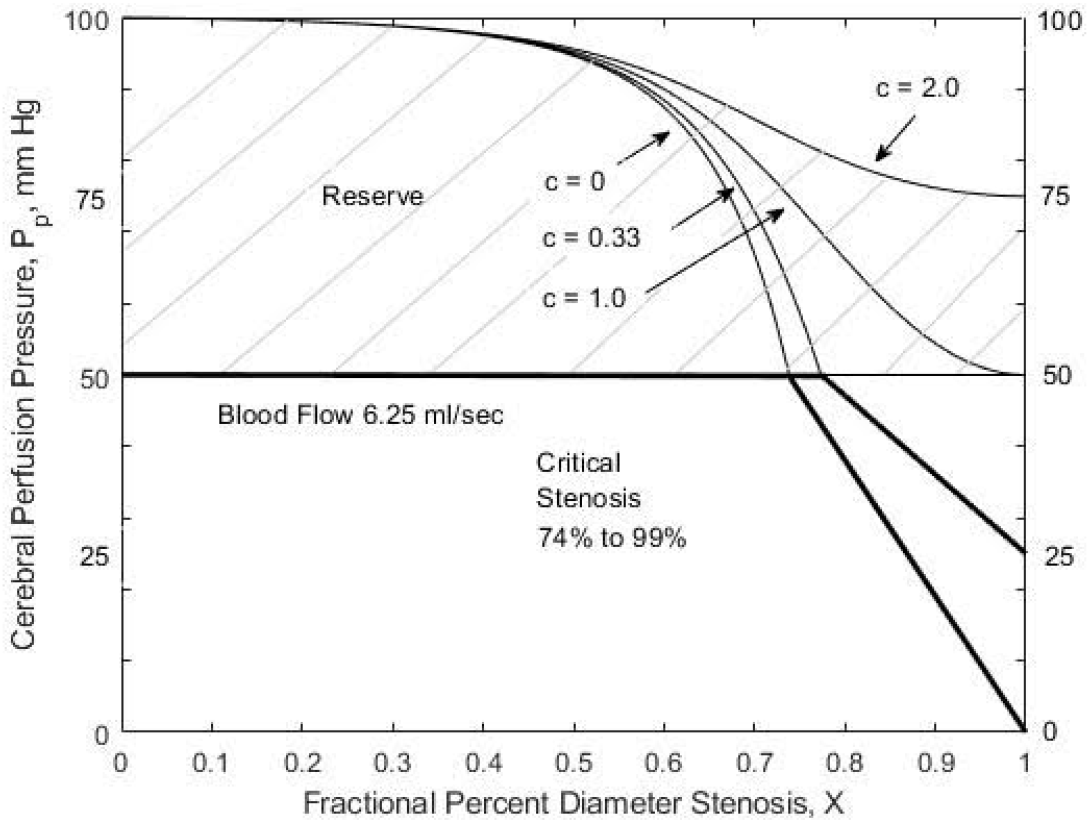
Numerical solution for cerebral hemisphere. Collateral blood flow potential determines critical stenosis values from 70% to 99% as collateral magnitude C increases from 0 to 1.0. The 74% is a typo. There is no critical stenosis when C > 1.0 and collaterals add to blood flow reserve. Blood flow heavy lines are in the lower flow-stenosis domain. As in Figures 3 and 4 the ischemic lines are slightly non-linear.

In the perfusion pressure zone P_p_ < P_cut_ blood flow is proportional to P_p_. Energy conservation is P_a_ = R_o_Q =(P_cut_/Q_n_)Q where Q = Q_s_ + Q_c_, Q < Q_n_ and the slope is Q_n_/P_cut_. When C > 0 but < 1.0, the solution for Q as a function of X is a solvable quadratic not included in Figure 5. In the absence of collateral blood flow (C = 0) Q is a near linear function of P_p_, similar to the Lassen model in Figure 1.

### Critical Arterial Stenosis

Critical arterial stenosis is defined as the percent diameter stenosis at which further stenosis progression reduces organ blood flow below normal into the ischemic zone. The system energy density equation P_a_ = (R_t_ + R_o_)Q. When Q = Q_n_ and R_o_ = P_cut_/Q_n_ and R_t_ = [(P_a_/Q_n_) – P_cut_/Q_n_] = (P_a_– P_cut_)/Q_n_. Because 1/R_t_ = 1/R_s_ + 1/R_c_, 1/Rt = Q_n_/(P_a_ – P_cut_) = [2A^2^/Q_s_K + KQ_n_/(P_a_ - P_cut_)]. Therefore, [Q_n_/(P_a_ - p_cut_)](1 - K) = [(2A^2^/Q_s_K) – 1]. Critical stenosis values occur when organ autoregulatory reserve energy dissipated arterial stenosis turbulent kinetic energy. For the three-component system; X_crit_ = 1 - 1/{[2A^2^(P_a_ - P_cut_)/Q_n_Q_s_(1 - C)] - 1}^1/4^. In addition, when P_p_ = P_cut_, Q_s_ = (1 – C)Q_n_ and the critical stenosis solution becomes X_crit_ = 1 – 1/{[2A^2^(P_a_ – P_cut_)/Q_n_^2^(1 – C)^2^] – 1}^1/4^. When C = 0, Rc = ∞, the absence of collateral blood flow, and X_crit_ = 1 - 1/{[2A^2^(P_a_ - P_cut_)/Q_n_^2^] - 1}^1/4^. If C = 1, X_crit_ = 1.0, 100% or occlusion. If C is higher than 1, X_crit_ does not exist because collateral blood flow is adequate to prevent ischemia, that is, collateral blood flow is adequate to maintain organ/tissue perfusion pressure, P_p_ higher than P_cut_.

Kidneys are a major solid organ with both closely autoregulated normal blood flow and essentially no collateral flow. Assuming renal artery blood flow 9.0 ml/sec, normal renal artery diameter 0.60cm, mean systemic arterial pressure 110mmHg and an autoregulation ischemic threshold pressure 60mmHg, renal artery critical stenosis is X_crit_ = 0.744 or 74.4% diameter stenosis. For a pressure threshold of 80mmHg for onset of increased renin production and glomerular filtration dysfunction, the critical diameter stenosis is 70.9%. However, glomerular flow does not contain blood cells. Glomerular dysfunction is more likely to have a critical stenosis value near 40% to 50%. This is illustrated in Figure 4.Critical stenosis values have a 95% confidence interval (CI) of approximately 3% for wide variance of normal anatomic, systemic arterial pressure and blood flow variables.

Cerebral hemispheres are major solid organs with closely autoregulated normal blood flow and highly variable between human collateral vascular resistance. In the absence of collateral blood flow and assuming normal internal carotid blood flow 6.25 ml/sec, normal diameter 0.50cm, mean systemic arterial pressure 100mmHg and autoregulation threshold 50mmHg, critical internal carotid artery (ICA) diameter stenosis is 0.704 or 70.4% with 3%CI. Based on clinically measurements^17^, mean internal carotid stump pressures is approximately 50 mmHg when normalized to P_a_ = 100mmHg, and cerebral collateral vascular resistance is close to hemisphere resistance, C = 1. For the approximate 50% of individuals with P_stump_ < 50mmHg (P_stump_ adjusted to P_a_ = 100mmHg) and collateral vascular resistance C < 1, (poor collaterals), critical ICA stenosis increases from 70% to 100% as C increases from 0 to 1.0. Conversely, if collateral blood flow is adequate to maintain normal cerebral flow with internal carotid occlusion, C > 1, (good collaterals), system critical stenosis does not exist because cerebral perfusion pressure is higher than critical (P_p_ > P_cut_) and reserve blood flow is available. This is illustrated in Figure 5.

### Arterial Stenosis Blood Flow

When collaterals are present, the energy relationship between collateral and arterial blood flow is Q_c_/Q_s_ = R_s_/R_c_ and Q = Q_c_ + Q_s_. For example, when organ/tissue perfusion pressure equal to or greater than threshold 50 mmHg, blood flow is normal and Q_c_ = Q_n_ -Q_s._ The result is Q_c_ = Q_s_^2^KQ_n_/2A^2^(P_a_ - P_cut_), a quadratic with solutions for Q_s_, and Q_c_. This is illustrated in Figure 6 when C = 1.0. For ICA diameter stenosis of 50%, 60%, 70%, 74% and 80% the corresponding ICA blood flows (Q_s_ and % of normal) are 5.80ml/sec (93%), 5.41 (87%), 4.45(71%), 3.85 (62%) and 2.74 (44%). Individuals with below average collateral blood flow, C < 1.0, have a lesser decrease in ICA blood flow. For ICA stump pressure of 25mmHg (C = 0.333) at 70% stenosis, Qs = 5.36 cm^3^/sec (86% of normal). Conversely, the approximate 50% of individuals with good collaterals (C > 1.0 and P_stump_ >50 mmHg) have a significant decrease in ICA blood flow. Similarly, these individuals will have much less energy dissipated in the stenosis zone (energy = P_p_Q_s_) than those with C < 1.0. Good collateral flow reduces both stenosis pressure gradient (P_a_ - P_p_) and ICA blood flow Q_s_ as stenosis progresses. For example, at 60% stenosis, individuals with average collaterals (C =1.0) have stenosis energy dissipation = (50mmHg)(5.4 cm ml/sec) = 270 energy units, versus those with good collaterals (35 mmHg)(4.3ml/sec) = 152 energy units.

**Figure 6.**
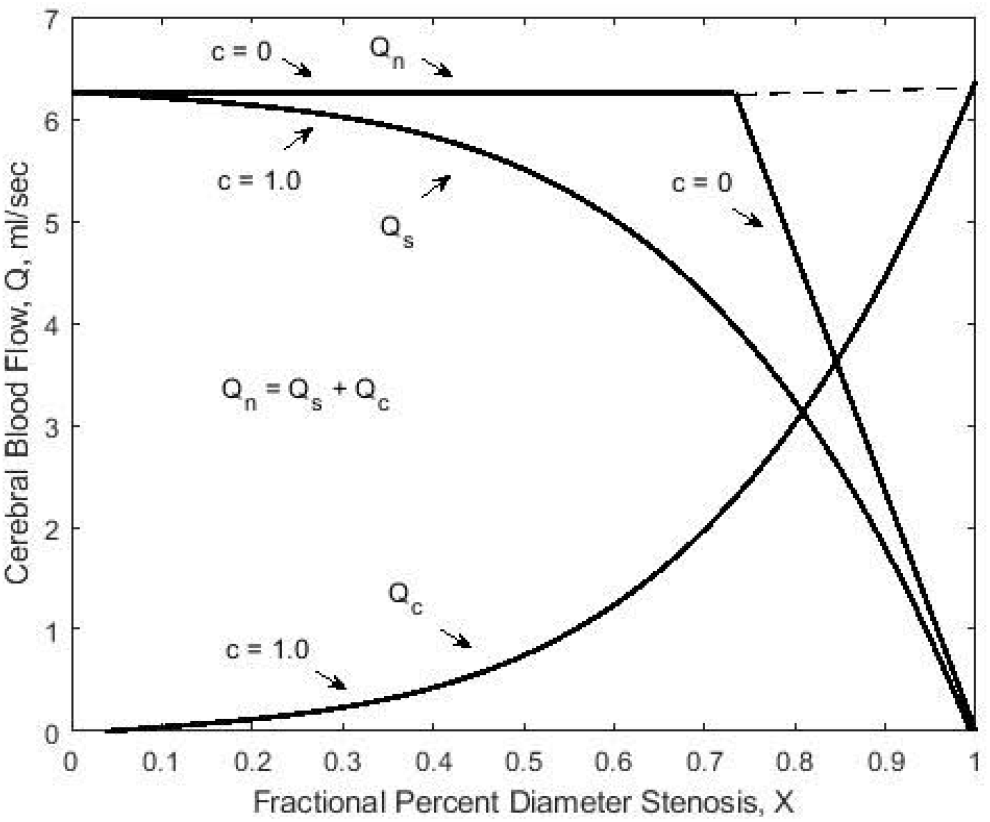
The two components of organ/tissue blood flow, stenotic artery, Q_s_, and collateral Q_c_, as a function of degree of stenosis when collateral magnitude C = 1.0. When C = 0, Q_c_ = 0.

## Discussion

Arterial stenosis reduces recipient organ/tissue’s perfusion pressure and blood flow reserve. If collaterals are absent, C = 0, there is a specific critical stenosis value. Renal artery ischemic critical stenosis is 74%, as in Figure 4. If collaterals are poor, C < 1.0, critical stenosis for body organ/tissues range from 65% to 99%, in proportion to collateral magnitude. For cerebral hemispheres this is 70% to 99%, as in Figure 5. If collaterals are good, C > 1.0, there are no critical values and reserve blood flow is available at occlusion, as also illustrated in Figure 5.

Turbulent vessel flow is an incompletely understood phenomena. Classical fluid mechanics is primarily based on momentum conservation that inadequately models arterial turbulence resulting in incomplete solutions, even with CFD. Conservation of energy requires stenosis TKE produce a pressure gradient. While unlikely to be original, the universal stenosis magnitude, K = {[1/(1-X)^4^ −1]}, where X is F%DS, is the energy dissipated by arterial stenosis. Of interest, the earlier proposed relationship between stenosis produced pressure gradients and stenosis area^4,5^ turns out to be area squared, diameter to the fourth power. The relationships betweenorgan/tissue perfusion pressure and blood flow as a function of X are illustrated in Figures 3, 4 and 5. Critical arterial stenosis only exists if the magnitude of collateral blood flow potential C is less than 1.0.

The defined magnitudes of stenosis energy, K, and collateral circulation, C, simplify presenting and understanding result. The K variable is specific for an organ/tissue system’s degree of artery stenosis. While K ranges from zero to infinity, the independent variable, X, ranges from zero to one, or 0% to 100% diameter stenosis. The C variable is specific between individuals and within an individual’s organ/tissue systems. Collateral circulation is a major determinant of the existence of ischemic critical stenosis. In the absence of collateral circulation C = 0, X_crit_ occurs when dissipated arterial stenosis energy negates autoregulation reserve energy, as for a renal artery stenosis. When collateral circulation is present with increasing C magnitude up to 1.0, (0 < C <1), X_crit_ increases up to 0.99, (99% stenosis). When collateral circulation is adequate to prevent organ/tissue autoregulation from being depleted and C > 1.0 if the artery occludes, blood flow is adequate to prevent ischemia and provide reserve.

### Potential Clinically Useful Results

Much of physical and biological science and resulting technology are based on models. They are considered good, but not necessarily correct if they are useful. A model is useful for both understanding and guidance for future investigation if it represents reality based on and consistent with clinical observation^18^. While this is a very simple model, the complex and somewhat difficult to understand results correlate with observed/measured clinical data and provide guidance for future investigation. Arterial stenosis generated kinetic energy magnitude, K, is appliable to any artery, as is stenosis vascular resistance. It can be easily modified for percent area stenosis allowing other stenosis geometries but not length. Collateral magnitude, C, is body organ/tissue system specific and individual specific for collateral blood flow potential. Critical arterial stenosis is a function of many variables including systemic arterial pressure. Hypertension may be beneficial in protecting organs/tissue with no or poor collaterals and arterial stenosis from ischemia. Common femoral/external iliac arteries likely have poor collaterals and may have critical stenosis values. Conversely, at rest an isolated superficial femoral artery stenosis has no critical value or produce ischemia, even with occlusion. However, exercise increases lower extremity metabolic energy requirements and collateral blood flow producing ischemic symptoms.

Progressive stenosis decreases artery blood flow, Q_s_, in proportion to the magnitude of collateral potential, as in Figure 6. Degree of arterial stenosis determined from ICA ultrasound velocities measurements may be underestimated, particularly if collateral flow is good. If degree of stenosis accurately measured by imaging is significantly higher than that estimated by ultrasound velocities, collateral flow is probably good. This may have some bearing on the important clinical problem of determining stroke risk and management recommendations for individuals with asymptomatic ICA stenosis. Good collateral blood flow potential is predicted to both prevent cerebral ischemia, even if the ICA occludes, and reduce stenosis zone energy dissipation. Further, the magnitude of turbulent energy dissipation may impact stenosis plaque stability. A decrease in ICA plaque zone TKE is potentially a further asset of good collaterals in individuals with asymptomatic carotid arterial stenosis.

### Limitations, Assumptions

While results appear complex, the analysis is relatively simple, algebraic without calculus or CFD. Blood flow and pressure are assumed to be steady, not pulsatile and out of phase. The validity of using turbulent kinetic energy produced by arterial stenosis to determine stenosis pressure gradient was confirmed by MRI for the aortic valve and phantom vessels^8-10^. While kinetic energy in normal arteries is laminar, mild stenosis of 30% - 50% diameter produce almost entirely turbulent kinetic energy^19,20^. Prior to these findings several attempt to describe arterial stenosis hemodynamics based on energy revert to a laminar flow momentum model or use empirical/experimental estimates to inadequately deal with turbulence^2,4,6^. Similar incomplete results were obtained using CFD turbulence models^3^.

Normal artery-collateral-organ/tissue systems have efficient laminar blood flow. The standard for estimating transition from laminar to turbulent flow is Reynolds number, Rn^11^. For steady flow in a pipe transition Rn is 2000 to 2500. Based on Womersley number^21^ and turbulent energy cascade transition^22^, vessel turbulent Rn are 400 to 500, close to the laminar Rn for renal and ICA examples. Depending on magnitude, stenosis dissipated TKE may clinically produce an audible bruit (frequency near 20 cycles/second) and/or a palpable thrill.

Mean systemic arterial pressure is normalized to include organ/tissue venous pressure. Numerical values for renal and ICA examples are established normal resting values for blood flow (mean velocities 30-36 cm/sec), arterial diameters and systemic pressure. The kidney autoregulation perfusion pressure thresholds of 60 mmHg for ischemia and 80mmHg for renin and glomerular filtration of 80 mmHg are established average values^23-26^. The cerebral hemisphere autoregulation lower threshold of 50mmHg is based on Lassen^14,15^.

## Conclusions

Critical arterial stenosis is a specific value for organs/tissues with no collateral flow. The magnitude of collateral blood flow potential C is C times normal organ/tissue flow Qn. Organs/tissues with poor collaterals (0<C<1) have a range of increasing critical diameter stenosis values with stenosis progression up to arterial occlusion depending on collateral magnitude. Organs/tissue systems with good collaterals (C>1) do not have critical stenosis. They have blood flow reserve that decreases with stenosis progression but remains available if the artery occludes.

## Data Availability

All data produced in the present work are contained in the manuscript

## Notes

Conflict of interest statement: This research did not receive any specific grant or funding agencies in the public, or non-profit sectors.

### Competing Interest Statement

The authors have declared no competing interest.

### Funding Statement

This study did not receive any funding

### Summary of Updates

Typos corrected

